# Impact of UTT on anticipated stigma among patients newly diagnosed with HIV in Johannesburg, South Africa

**DOI:** 10.1101/2025.05.28.25328494

**Authors:** Tembeka Sineke, Idah Mokhele, Robert AC Ruiter, Dorina Onoya, Mandisa Dukashe

## Abstract

**Background:** Anticipated stigma – the fear that HIV diagnosis and status disclosure could have negative social implications – may adversely affect engagement with HIV care and treatment, despite universal eligibility for treatment under universal-test-and-treat (UTT). We aimed to determine the prevalence and predictors of anticipated stigma among newly HIV-diagnosed individuals in the context of universal access to treatment in Johannesburg, South Africa.

**Methods:** We conducted a cross-sectional survey of 652 newly HIV-diagnosed adults (≥18 years) (64.1% female, with a median age of 33 years and an interquartile range [IQR] of 28–39 years) enrolled from October 2017 to August 2018 at four primary healthcare clinics in Johannesburg. Participants were interviewed immediately after receiving their HIV test results. We used an adapted five-item, four-point scale measuring agreement with statements regarding HIV disclosure concerns and HIV status concealment (Cronbach’s alpha =.82). Mean scores were categorized as “low-to-medium” (score<=2.5), or “high” (score>2.5). We used Modified Poisson regression to assess predictors of high anticipated stigma and report adjusted risk ratios (aRR) with 95% confidence intervals (CIs).

**Results:** Overall, 55% of study participants had high anticipated stigma; 55.8% for males, 61.1% for 18–29-year-olds, and 43% for those married. Individuals in an unmarried relationship were more likely to experience high anticipated stigma than those married (aRR 1.10, 95% CI: 1.01-1.18). High anticipated stigma was lower among: older individuals (aRR 0.94 for being 30-39 vs 18-29 years, 95% CI: 0.88-0.99), those with a primary home in another province/rural area (aRR 0.82 another province vs current house, 95% CI: 0.78-0.87) or another country (aRR 0.83 another country vs current house, 95% CI: 0.78-0.88), those living in current homes for ≥5 years (aRR 0.93 for >5 years vs <1 year, 95% CI: 0.88-0.99), those with low ART concerns (aRR 0.86, 95 % CI: 0.82-0.90), and those with low perceived social-support (aRR 0.79 for low vs high, 95 % CI: 0.70-0.88).

**Conclusion:** Over 50% of adults diagnosed with HIV in the UTT era had high anticipated stigma. Findings highlight the need to address factors that continue to drive anticipated stigma, to improve social integration and mitigate the potential impact on engagement in HIV care. In addition, enhancing coping skills among individuals living with HIV is crucial.

## INTRODUCTION

South Africa has made substantial progress towards achieving the 95-95-95 targets set by UNAIDS[1], reaching 95.4% of people living with HIV (PLHIV) diagnosed, of those diagnosed, 78.7% on Antiretroviral Treatment (ART), and 91.3% of those on treatment achieving viral suppression as of 2023[2]. In recent years, access to ART is widespread, and HIV has become more of a chronic health condition as people living with HIV (PLHIV) can now live longer. However, while the same-day ART initiation has been an essential step in increasing accessibility[3, 4], it does not directly address social challenges that PLHIV continue to face[5]. While treatment improves the personal health and longevity outlook of the individual living with HIV, it does not address the social reintegration needed to facilitate the return to normal social interactions.

HIV denialism previously fuelled stigma, questioning the link between HIV and AIDS, and delaying the rollout of ART[6]. This was coupled with misinformation, shame, and reluctance to disclose one’s status[7]. The stigma index has significantly reduced over time, particularly external and social stigma in South Africa, which includes denial of some services for PLHIV; however, internalised stigma, characterised by feelings of low self-worth, guilt, and shame, persists[8]. This internal stigma fuels anticipated social stigma, resulting in self-isolation that has a detrimental impact on PLHIV’s quality of life. Recent work has shown that PLHIV still anticipate and experience stigma in the context of romantic relationships (starting or maintaining)[9].

While ART access has improved in South Africa, treatment disengagement and inconsistent adherence to ART remain a threat to achieving sustained viral suppression and eliminating HIV in resource-limited settings[10–12]. Anticipated stigma is defined as a belief and expectation by PLHIV of future repercussions and ill treatment due to their HIV positive status. PLHIV who anticipate stigma may withdraw from social relationships in an attempt to minimise potential discrimination, leading to social isolation and withdrawal from potentially supportive networks[13]. Among other factors, anticipated stigma is a significant predictor of poor adherence [14–16]. Anticipated stigma has also been associated with distrust of healthcare workers [17], increased medication concerns, and low treatment knowledge. PLHIV also fear that engaging in ART care may unintentionally reveal one’s HIV status through regular clinic visits and carrying medication or printed medical information[13]. PLHIV have voiced concerns that unintentional disclosure would result in stigmatisation[18].

Other studies have shown that anticipated stigma negatively impacts testing for HIV, a barrier to ART initiation, and several studies have shown HIV-related stigma to be associated with depressive symptoms among PLHIV[19, 20]. This may result in social withdrawal and isolation as individuals tend to suppress their emotions due to fears of being stigmatised[21].

The National Strategic Plan (NSP) for 2023 identified stigma as one of the important issues to be addressed. It called for interventions to address this[22]. These include evidence-based interventions, community engagement, and collaborations between different sectors. In light of the changes in treatment guidelines, we aimed to determine the prevalence and predictors of anticipated stigma among newly HIV-diagnosed individuals under the UTT policy in Johannesburg, South Africa.

## MATERIALS AND METHODS

The enrolment of patients was conducted according to previously described criteria [23]. Patients were interviewed immediately after HIV diagnosis, and ART initiation was determined through medical record review up to six months post-test. Participants self-reported being newly diagnosed during the screening process. Enrolled patients with a prior history of ART were excluded from the analytic dataset. We also excluded patients who were psychologically unable or too sick to participate, unwilling to provide consent or planned to get treatment elsewhere. Additionally, women who were pregnant at HIV diagnosis were excluded from the study because in-pregnancy treatment initiation and care processes differ from those of non-pregnant women. Our study population included participants with a median age of 33 years (IQR: 28.0-39.0), 64.1% were female, 58.9% had some secondary level of education and 14.1% were married.

### Variable definitions

We used an adapted five-item, four-point scale (1-“strongly agree” to 5-“strongly disagree”) measuring agreement with statements regarding HIV disclosure concerns and HIV status concealment. Questions included concerns regarding rejection, fear of being judged, concerns about the risk of disclosure to others, feelings of shame regarding the diagnosis, and a need to keep the diagnosis as a secret. All questions were closed-ended with options For the final analysis, we dichotomized the final outcome of anticipated stigma whereby mean scores were categorized as “low-to-medium” (if the score<=2.5), or “high” (if the score>2.5)[24, 25].

Other socio-demographic factors assessed included age, sex, highest education completed, marital status, employment status, whether the patient was the household breadwinner, the number of child dependents the patient had, and primary source of income. Factors related to health care access, including the history of visiting the testing clinic or any other health provider, and HIV testing history were also assessed. Additional factors also included a history of sexual risk behaviour, including condom usage at last sex, number of sexual partners in the previous twelve months and to whom the patients had disclosed their intention to come for HIV testing, and whether the patients were accompanied by anyone to the testing clinic[23].

The study protocol was reviewed and approved by the Human Research Ethics Committee of the University of Witwatersrand (M1704122). All personal identifiers were removed from the final analytic dataset.

### Statistical analysis

We describe the characteristics of study participants using proportions, frequencies, means with standard deviation (SD), and medians with interquartile ranges (IQRs) as appropriate. Factors associated with the prevalence of high anticipated stigma among newly diagnosed patients were assessed using the Modified Poisson regression and report adjusted risk ratios (aRR) with 95% confidence intervals (CIs). Data analysis was conducted using STATA version 14 (StataCorp, College Station, TX).

## RESULTS

### Demographic and clinical characteristics of newly diagnosed HIV patients by their gender

A total of 652 newly HIV diagnosed participants were enrolled (**Table 1**). Participants had a median age of 33 years (IQR: 28.0-39.0), 64.1% were female, and 35.3% were married. Over half of the participants (51.5%) spoke Nguni languages (which include: Zulu, Xhosa, Ndebele, Swati, etc.). More than half of the participants were employed and 47.2% of the employed were female. Additionally, only 62.4% had disclosed their intention to test for HIV before the clinic visit and 26.0% were accompanied to the testing site. However, participants’ baseline intention to start ART was nearly universal at 99.4%, with only 0.6% planning to never start ART. Similarly, 96.3% of participants intended to disclose their HIV-positive status, 4.7% had low perceived social support, and 67% of those were females, and 52% had medium concerns about ART.

**Table 1.**
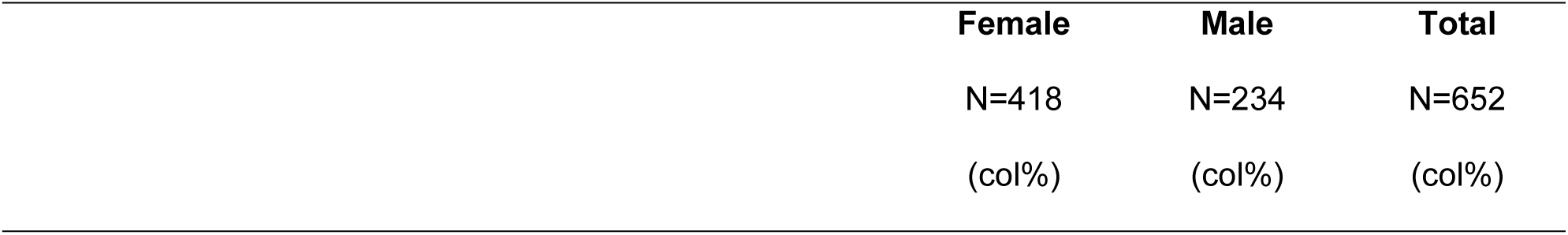
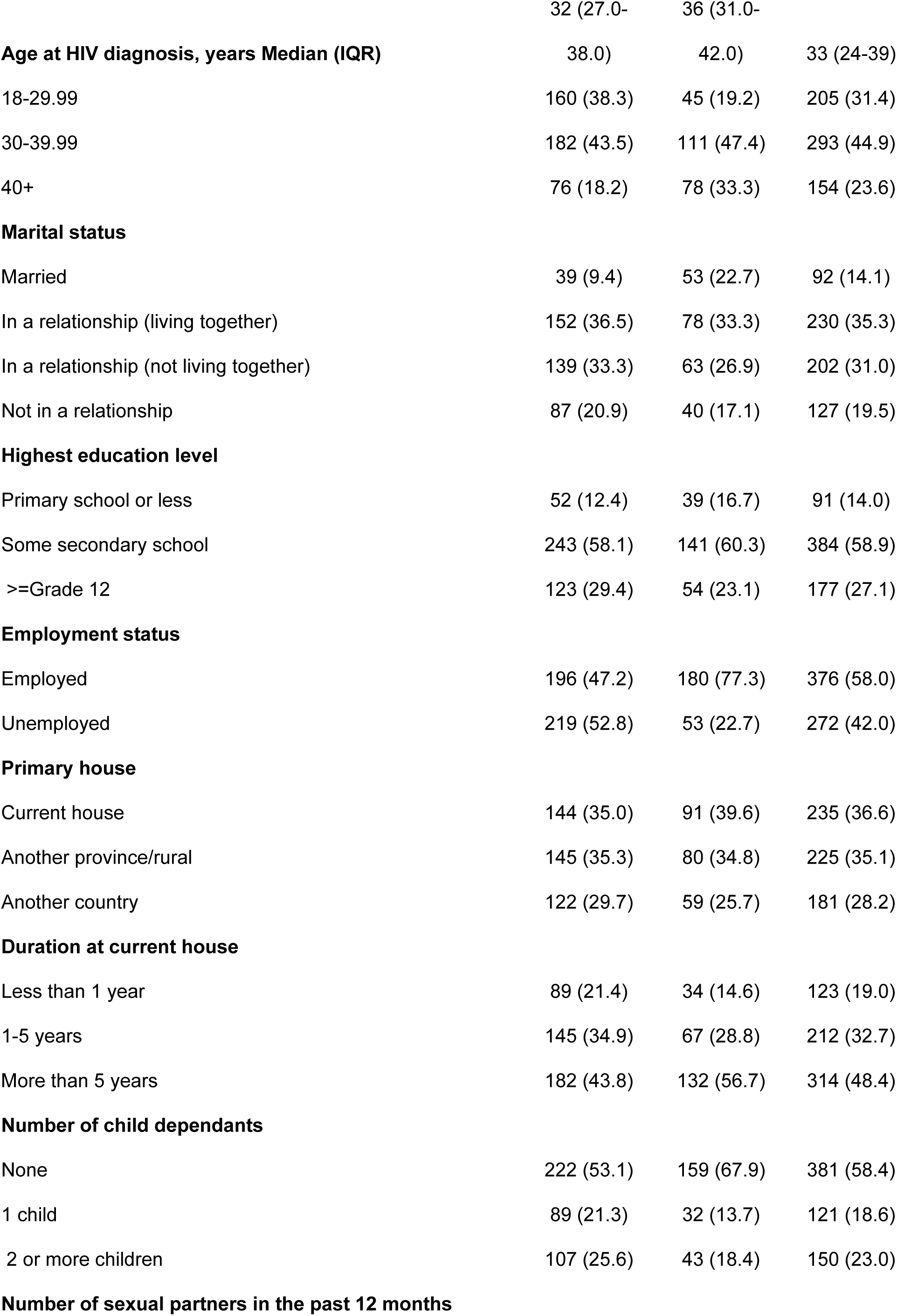
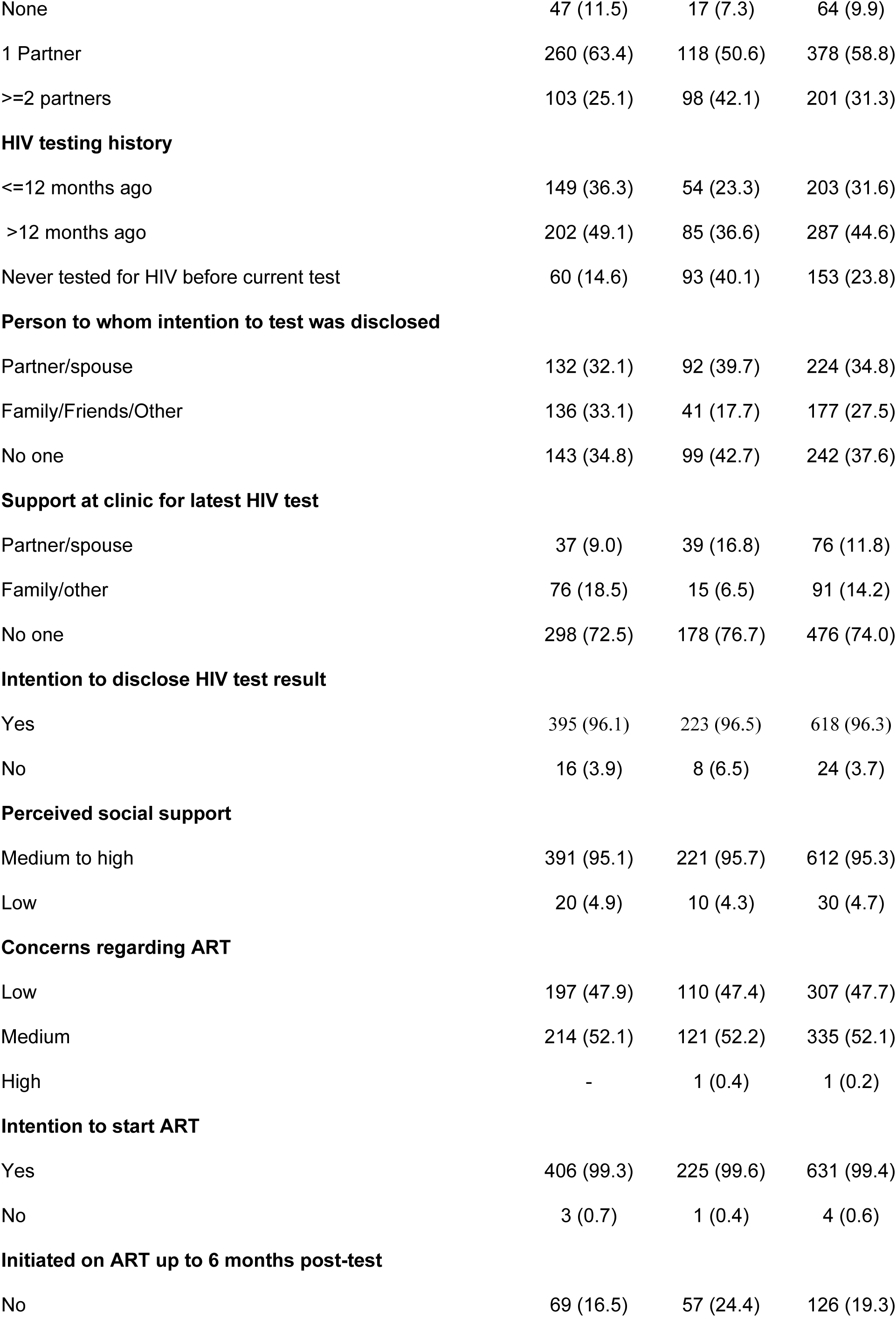
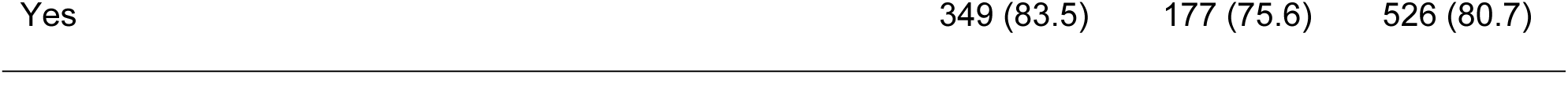
Participant sociodemographic characteristics (n=652)

|  | **Female** | **Male** | **Total** |
| --- | --- | --- | --- |
|  | N=418 | N=234 | N=652 |
|  | (col%) | (col%) | (col%) |
|  | 32 (27.0- | 36 (31.0- |  |
| **Age at HIV diagnosis, years Median (IQR)** | 38.0) | 42.0) | 33 (24-39) |
| 18-29.99 | 160 (38.3) | 45 (19.2) | 205 (31.4) |
| 30-39.99 | 182 (43.5) | 111 (47.4) | 293 (44.9) |
| 40+ | 76 (18.2) | 78 (33.3) | 154 (23.6) |
| **Marital status** |  |  |  |
| Married | 39 (9.4) | 53 (22.7) | 92 (14.1) |
| In a relationship (living together) | 152 (36.5) | 78 (33.3) | 230 (35.3) |
| In a relationship (not living together) | 139 (33.3) | 63 (26.9) | 202 (31.0) |
| Not in a relationship | 87 (20.9) | 40 (17.1) | 127 (19.5) |
| **Highest education level** |  |  |  |
| Primary school or less | 52 (12.4) | 39 (16.7) | 91 (14.0) |
| Some secondary school | 243 (58.1) | 141 (60.3) | 384 (58.9) |
| >=Grade 12 | 123 (29.4) | 54 (23.1) | 177 (27.1) |
| **Employment status** |  |  |  |
| Employed | 196 (47.2) | 180 (77.3) | 376 (58.0) |
| Unemployed | 219 (52.8) | 53 (22.7) | 272 (42.0) |
| **Primary house** |  |  |  |
| Current house | 144 (35.0) | 91 (39.6) | 235 (36.6) |
| Another province/rural | 145 (35.3) | 80 (34.8) | 225 (35.1) |
| Another country | 122 (29.7) | 59 (25.7) | 181 (28.2) |
| **Duration at current house** |  |  |  |
| Less than 1 year | 89 (21.4) | 34 (14.6) | 123 (19.0) |
| 1-5 years | 145 (34.9) | 67 (28.8) | 212 (32.7) |
| More than 5 years | 182 (43.8) | 132 (56.7) | 314 (48.4) |
| **Number of child dependants** |  |  |  |
| None | 222 (53.1) | 159 (67.9) | 381 (58.4) |
| 1 child | 89 (21.3) | 32 (13.7) | 121 (18.6) |
| 2 or more children | 107 (25.6) | 43 (18.4) | 150 (23.0) |
| **Number of sexual partners in the past 12 months** |  |  |  |
| None | 47 (11.5) | 17 (7.3) | 64 (9.9) |
| 1 Partner | 260 (63.4) | 118 (50.6) | 378 (58.8) |
| >=2 partners | 103 (25.1) | 98 (42.1) | 201 (31.3) |
| **HIV testing history** |  |  |  |
| <=12 months ago | 149 (36.3) | 54 (23.3) | 203 (31.6) |
| >12 months ago | 202 (49.1) | 85 (36.6) | 287 (44.6) |
| Never tested for HIV before current test | 60 (14.6) | 93 (40.1) | 153 (23.8) |
| **Person to whom intention to test was disclosed** |  |  |  |
| Partner/spouse | 132 (32.1) | 92 (39.7) | 224 (34.8) |
| Family/Friends/Other | 136 (33.1) | 41 (17.7) | 177 (27.5) |
| No one | 143 (34.8) | 99 (42.7) | 242 (37.6) |
| **Support at clinic for latest HIV test** |  |  |  |
| Partner/spouse | 37 (9.0) | 39 (16.8) | 76 (11.8) |
| Family/other | 76 (18.5) | 15 (6.5) | 91 (14.2) |
| No one | 298 (72.5) | 178 (76.7) | 476 (74.0) |
| **Intention to disclose HIV test result** |  |  |  |
| Yes | 395 (96.1) | 223 (96.5) | 618 (96.3) |
| No | 16 (3.9) | 8 (6.5) | 24 (3.7) |
| **Perceived social support** |  |  |  |
| Medium to high | 391 (95.1) | 221 (95.7) | 612 (95.3) |
| Low | 20 (4.9) | 10 (4.3) | 30 (4.7) |
| **Concerns regarding ART** |  |  |  |
| Low | 197 (47.9) | 110 (47.4) | 307 (47.7) |
| Medium | 214 (52.1) | 121 (52.2) | 335 (52.1) |
| High | - | 1 (0.4) | 1 (0.2) |
| **Intention to start ART** |  |  |  |
| Yes | 406 (99.3) | 225 (99.6) | 631 (99.4) |
| No | 3 (0.7) | 1 (0.4) | 4 (0.6) |
| **Initiated on ART up to 6 months post-test** |  |  |  |
| No | 69 (16.5) | 57 (24.4) | 126 (19.3) |
| Yes | 349 (83.5) | 177 (75.6) | 526 (80.7) |

**Table 2.**
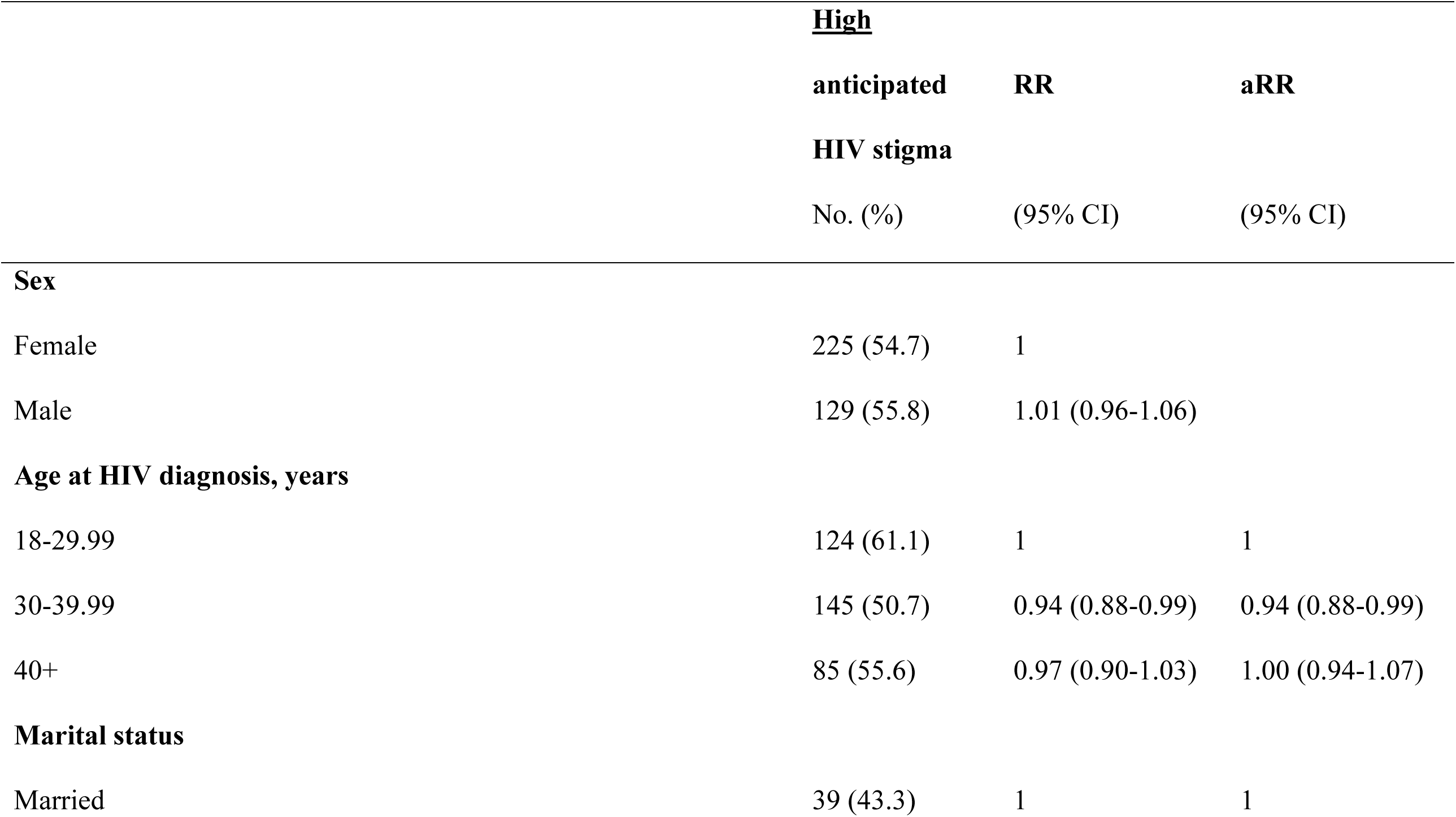
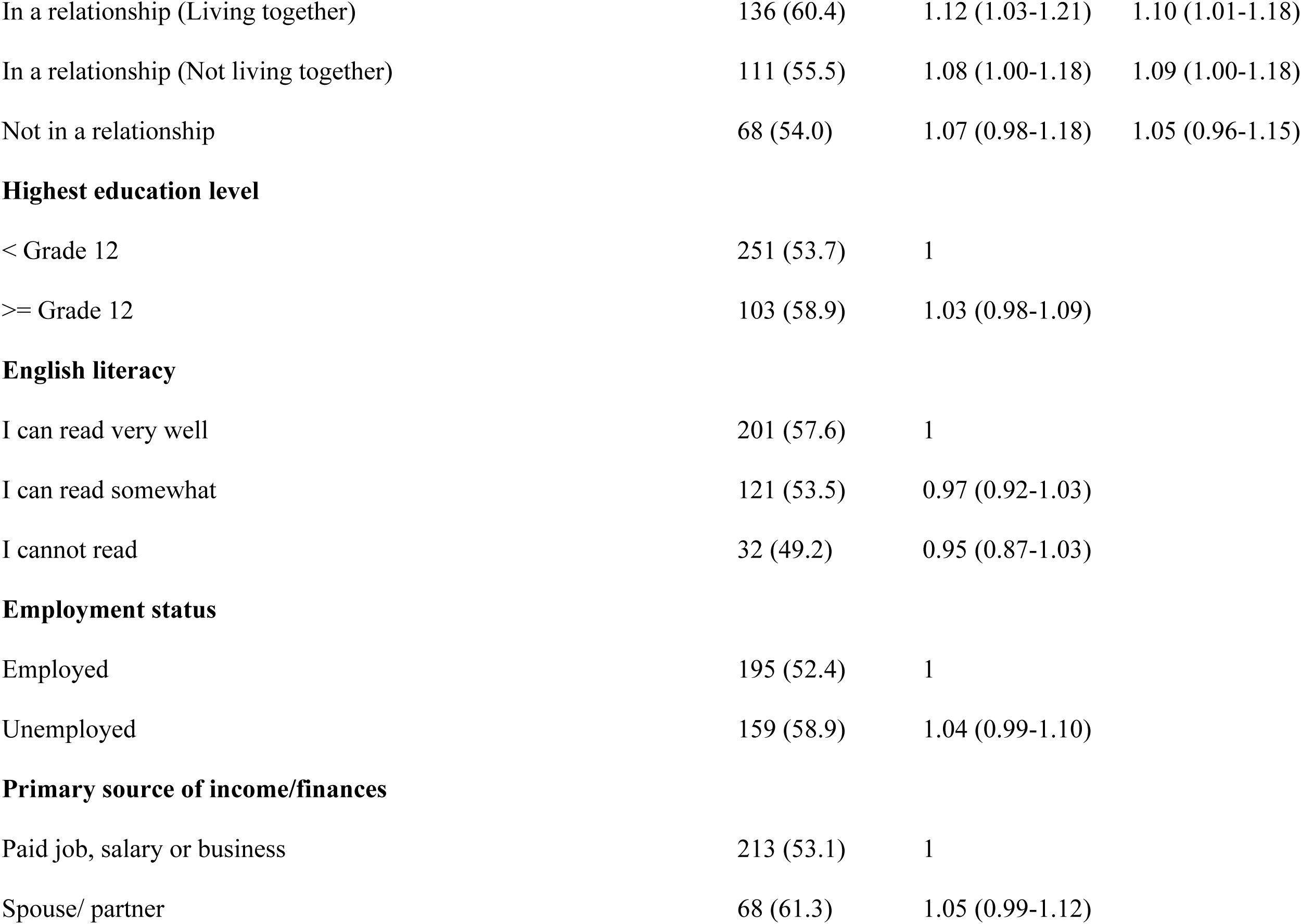
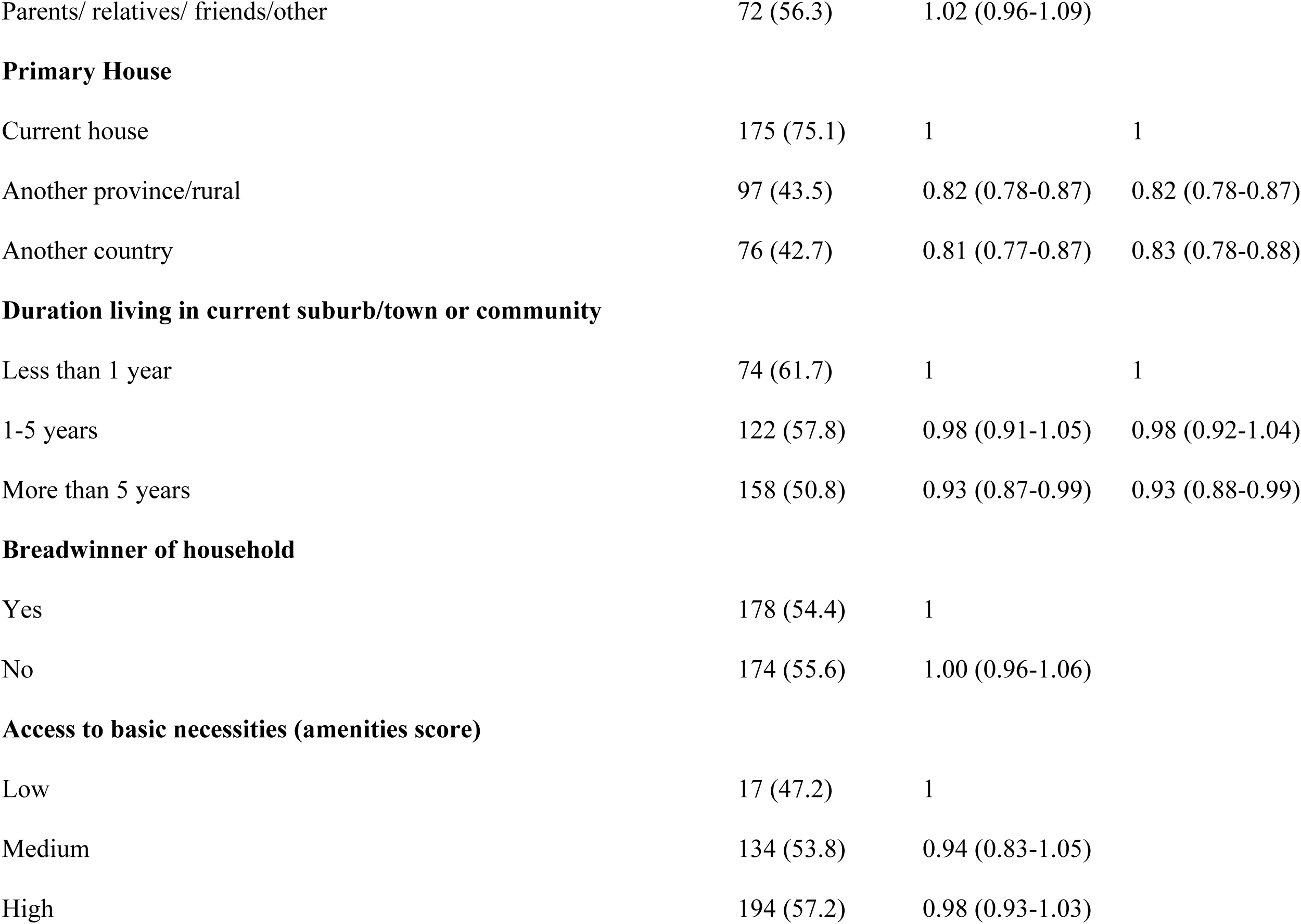
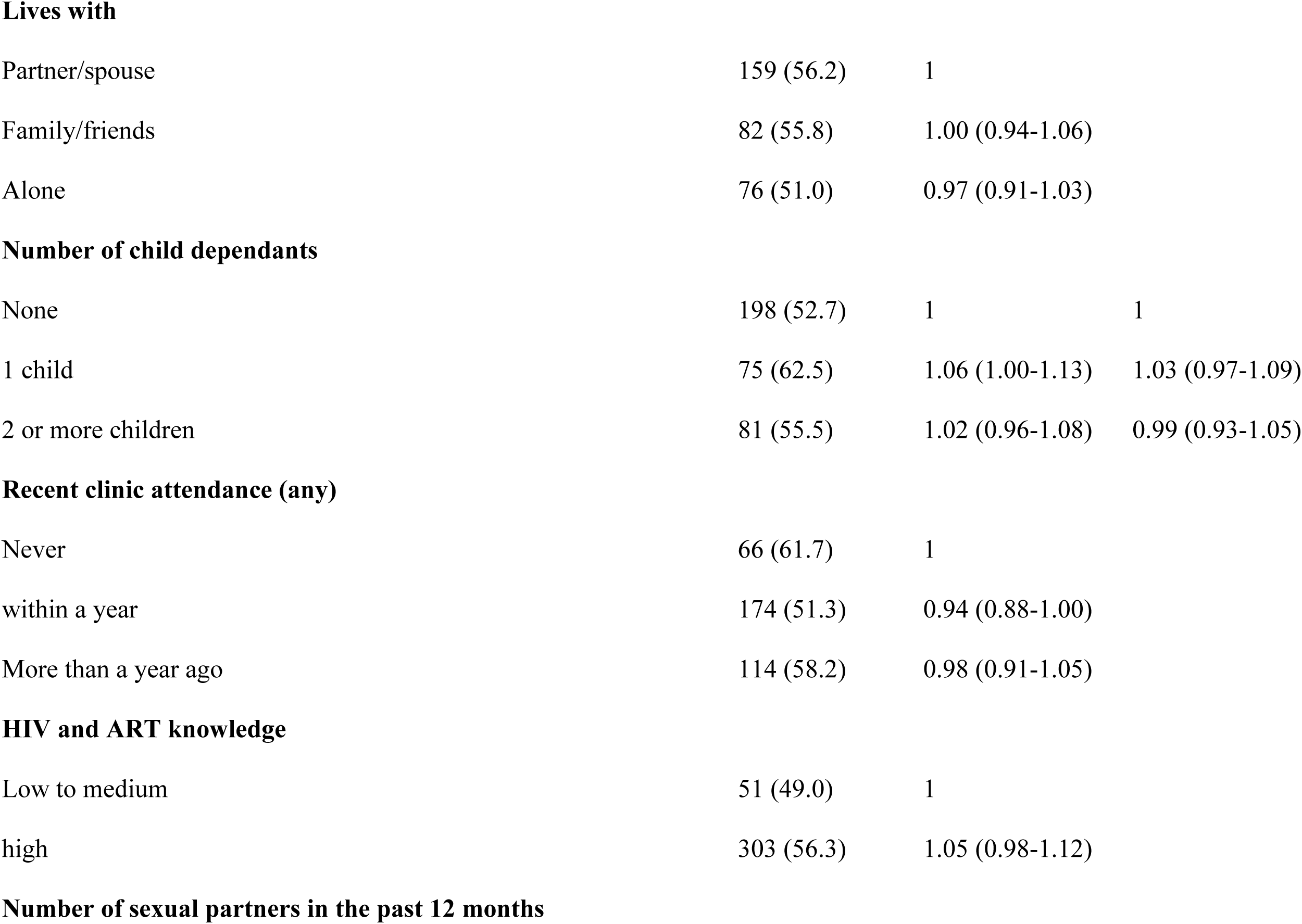
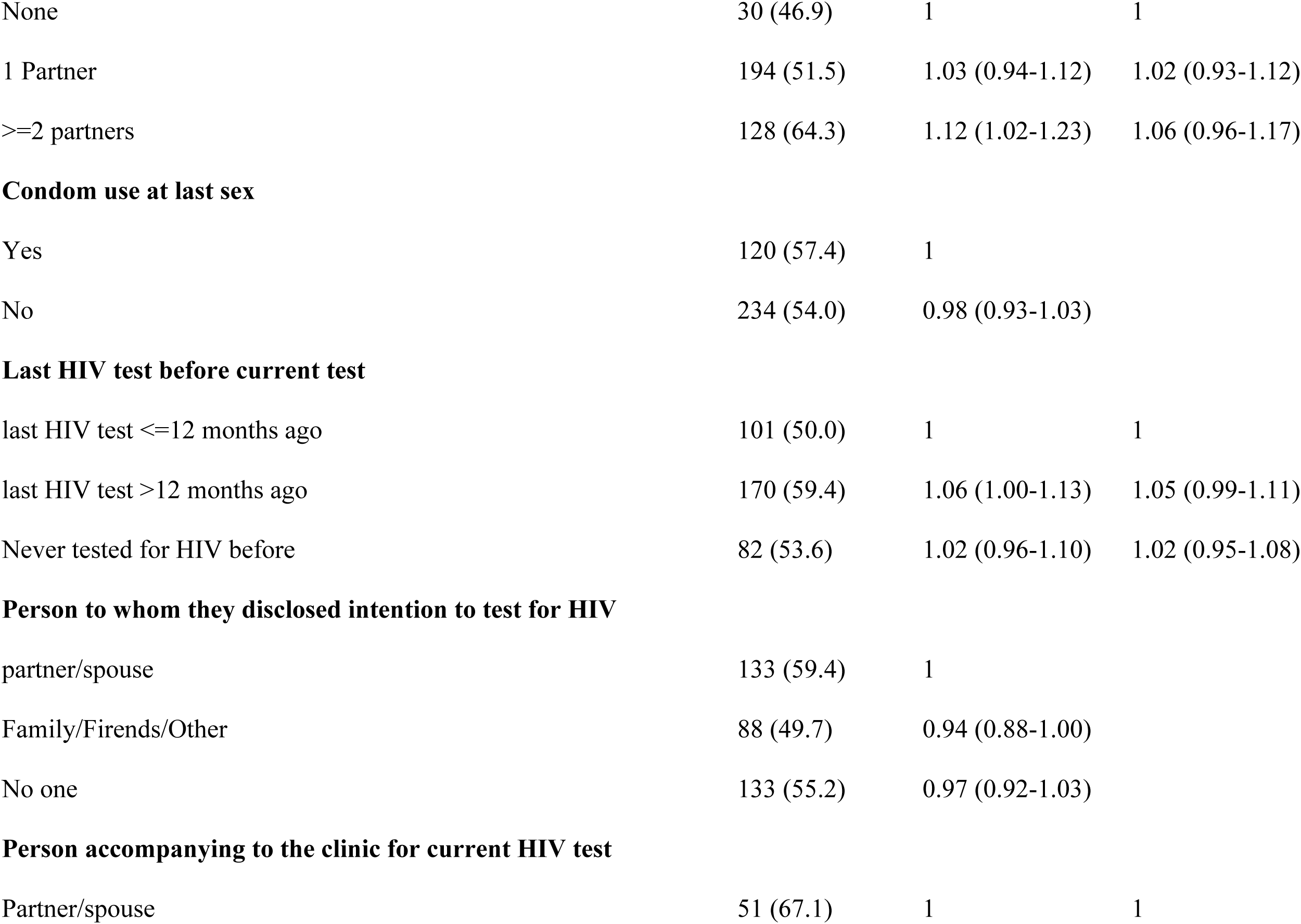
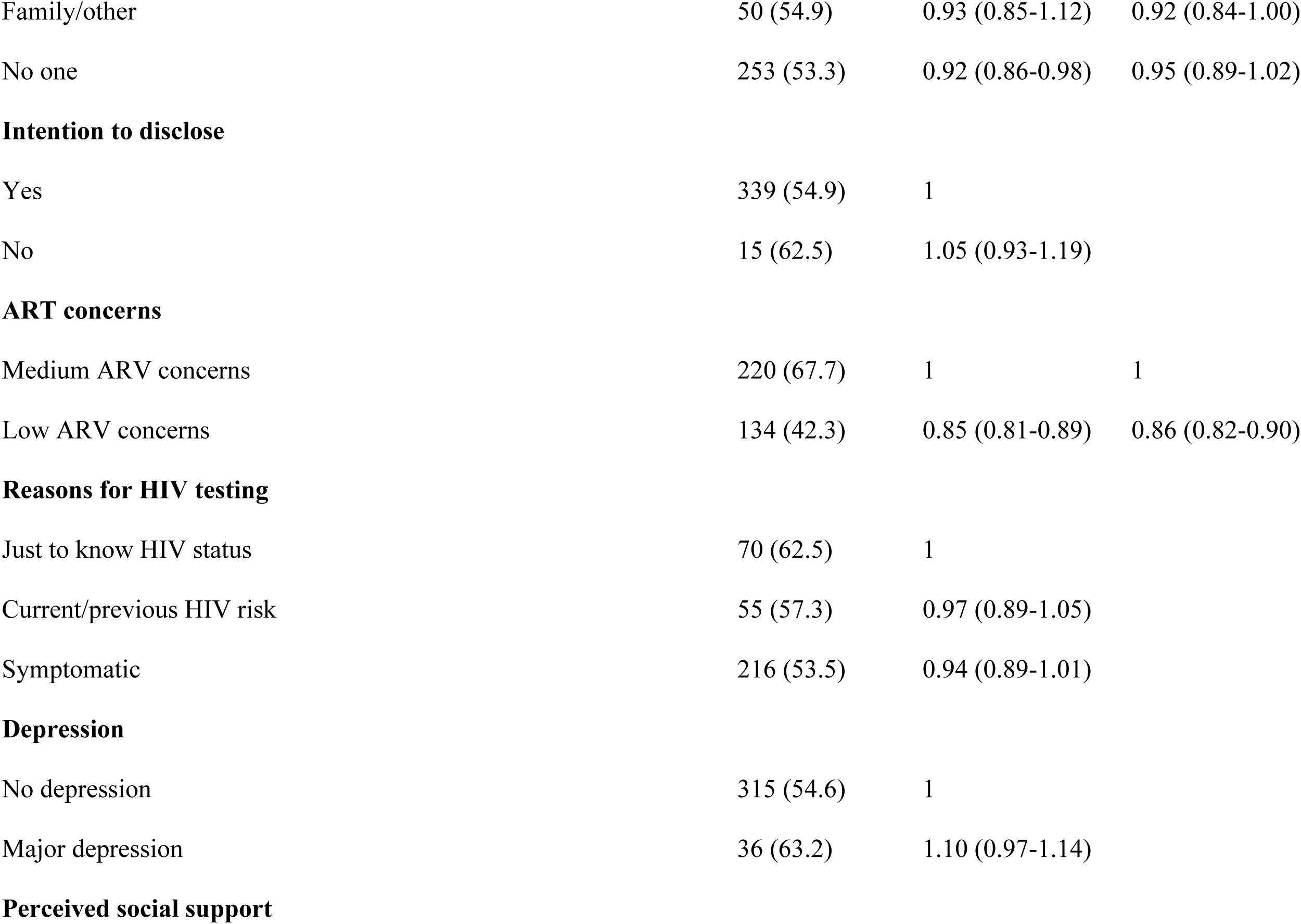
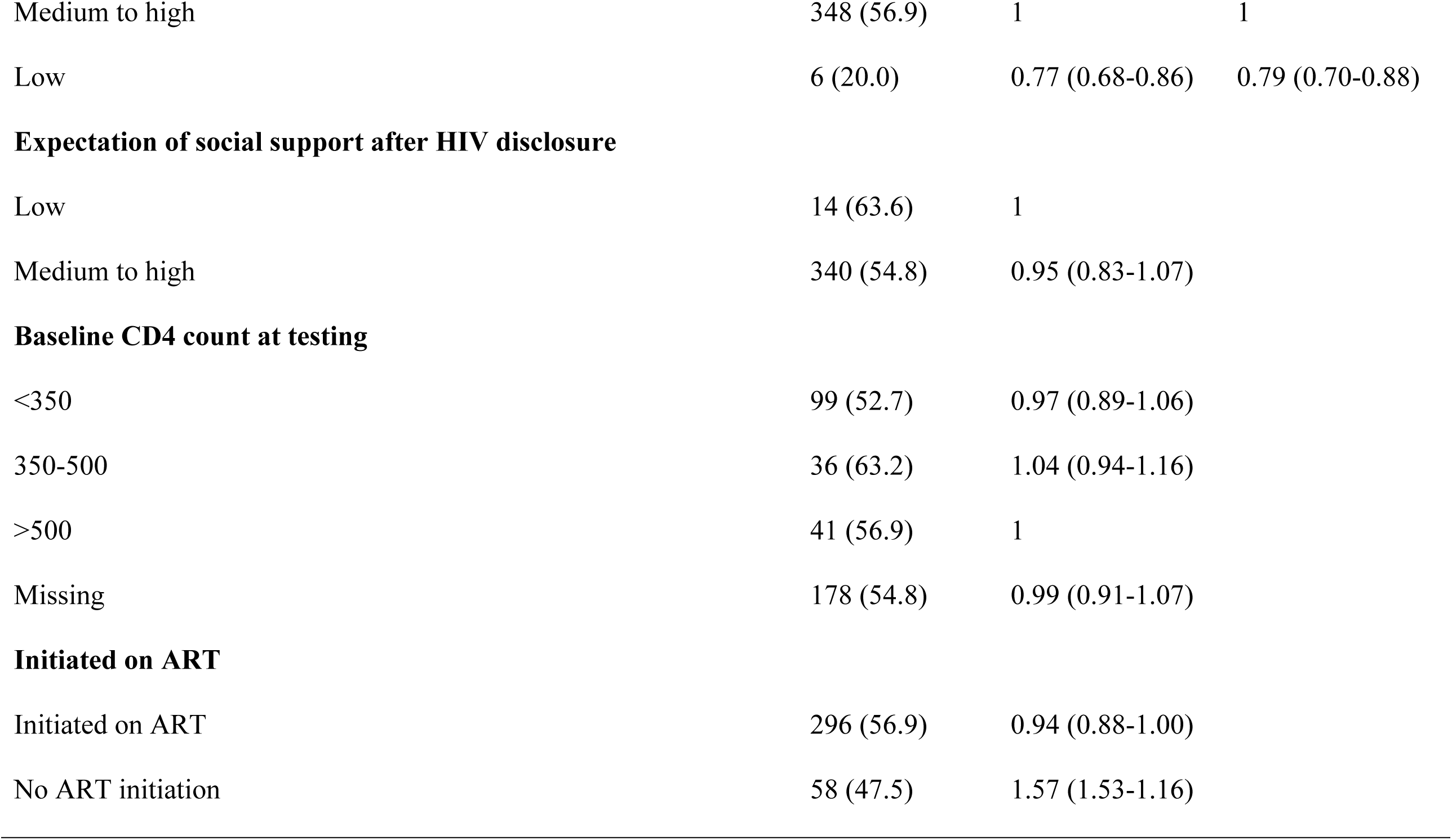
Correlates of high anticipated HIV stigma among newly diagnosed.

|  | <u>High</u> |  |  |
| --- | --- | --- | --- |
|  | anticipated | RR | aRR |
|  | HIV stigma |  |  |
|  | No. (%) | (95% CI) | (95% CI) |
| **Sex** |  |  |  |
| Female | 225 (54.7) | 1 |  |
| Male | 129 (55.8) | 1.01 (0.96-1.06) |  |
| **Age at HIV diagnosis, years** |  |  |  |
| 18-29.99 | 124 (61.1) | 1 | 1 |
| 30-39.99 | 145 (50.7) | 0.94 (0.88-0.99) | 0.94 (0.88-0.99) |
| 40+ | 85 (55.6) | 0.97 (0.90-1.03) | 1.00 (0.94-1.07) |
| **Marital status** |  |  |  |
| Married | 39 (43.3) | 1 | 1 |
| In a relationship (Living together) | 136 (60.4) | 1.12 (1.03-1.21) | 1.10 (1.01-1.18) |
| In a relationship (Not living together) | 111 (55.5) | 1.08 (1.00-1.18) | 1.09 (1.00-1.18) |
| Not in a relationship | 68 (54.0) | 1.07 (0.98-1.18) | 1.05 (0.96-1.15) |

**Highest education level**
|  |  |  |
| --- | --- | --- |
| < Grade 12 | 251 (53.7) | 1 |
| >= Grade 12 | 103 (58.9) | 1.03 (0.98-1.09) |

**English literacy**
|  |  |  |
| --- | --- | --- |
| I can read very well | 201 (57.6) | 1 |
| I can read somewhat | 121 (53.5) | 0.97 (0.92-1.03) |
| I cannot read | 32 (49.2) | 0.95 (0.87-1.03) |

**Employment status**
|  |  |  |
| --- | --- | --- |
| Employed | 195 (52.4) | 1 |
| Unemployed | 159 (58.9) | 1.04 (0.99-1.10) |
| **Primary source of income/finances** |  |  |  |
| Paid job, salary or business | 213 (53.1) | 1 |  |
| Spouse/ partner | 68 (61.3) | 1.05 (0.99-1.12) |  |
| Parents/ relatives/ friends/other | 72 (56.3) | 1.02 (0.96-1.09) |  |
| **Primary House** |  |  |  |
| Current house | 175 (75.1) | 1 | 1 |
| Another province/rural | 97 (43.5) | 0.82 (0.78-0.87) | 0.82 (0.78-0.87) |
| Another country | 76 (42.7) | 0.81 (0.77-0.87) | 0.83 (0.78-0.88) |
| **Duration living in current suburb/town or community** |  |  |  |
| Less than 1 year | 74 (61.7) | 1 | 1 |
| 1-5 years | 122 (57.8) | 0.98 (0.91-1.05) | 0.98 (0.92-1.04) |
| More than 5 years | 158 (50.8) | 0.93 (0.87-0.99) | 0.93 (0.88-0.99) |
| **Breadwinner of household** |  |  |  |
| Yes | 178 (54.4) | 1 |  |
| No | 174 (55.6) | 1.00 (0.96-1.06) |  |
| **Access to basic necessities (amenities score)** |  |  |  |
| Low | 17 (47.2) | 1 |  |
| Medium | 134 (53.8) | 0.94 (0.83-1.05) |  |
| High | 194 (57.2) | 0.98 (0.93-1.03) |  |

**Lives with**
|  |  |  |
| --- | --- | --- |
| Partner/spouse | 159 (56.2) | 1 |
| Family/friends | 82 (55.8) | 1.00 (0.94-1.06) |
| Alone | 76 (51.0) | 0.97 (0.91-1.03) |
| **Number of child dependants** |  |  |  |
| None | 198 (52.7) | 1 | 1 |
| 1 child | 75 (62.5) | 1.06 (1.00-1.13) | 1.03 (0.97-1.09) |
| 2 or more children | 81 (55.5) | 1.02 (0.96-1.08) | 0.99 (0.93-1.05) |

**Recent clinic attendance (any)**
|  |  |  |
| --- | --- | --- |
| Never | 66 (61.7) | 1 |
| within a year | 174 (51.3) | 0.94 (0.88-1.00) |
| More than a year ago | 114 (58.2) | 0.98 (0.91-1.05) |

**HIV and ART knowledge**
|  |  |  |
| --- | --- | --- |
| Low to medium | 51 (49.0) | 1 |
| high | 303 (56.3) | 1.05 (0.98-1.12) |
| **Number of sexual partners in the past 12 months** |  |  |  |
| None | 30 (46.9) | 1 | 1 |
| 1 Partner | 194 (51.5) | 1.03 (0.94-1.12) | 1.02 (0.93-1.12) |
| >=2 partners | 128 (64.3) | 1.12 (1.02-1.23) | 1.06 (0.96-1.17) |
| **Condom use at last sex** |  |  |  |
| Yes | 120 (57.4) | 1 |  |
| No | 234 (54.0) | 0.98 (0.93-1.03) |  |
| **Last HIV test before current test** |  |  |  |
| last HIV test <=12 months ago | 101 (50.0) | 1 | 1 |
| last HIV test >12 months ago | 170 (59.4) | 1.06 (1.00-1.13) | 1.05 (0.99-1.11) |
| Never tested for HIV before | 82 (53.6) | 1.02 (0.96-1.10) | 1.02 (0.95-1.08) |
| **Person to whom they disclosed intention to test for HIV** |  |  |  |
| partner/spouse | 133 (59.4) | 1 |  |
| Family/Firends/Other | 88 (49.7) | 0.94 (0.88-1.00) |  |
| No one | 133 (55.2) | 0.97 (0.92-1.03) |  |
| **Person accompanying to the clinic for current HIV test** |  |  |  |
| Partner/spouse | 51 (67.1) | 1 | 1 |
| Family/other | 50 (54.9) | 0.93 (0.85-1.12) | 0.92 (0.84-1.00) |
| No one | 253 (53.3) | 0.92 (0.86-0.98) | 0.95 (0.89-1.02) |
| **Intention to disclose** |  |  |  |
| Yes | 339 (54.9) | 1 |  |
| No | 15 (62.5) | 1.05 (0.93-1.19) |  |
| **ART concerns** |  |  |  |
| Medium ARV concerns | 220 (67.7) | 1 | 1 |
| Low ARV concerns | 134 (42.3) | 0.85 (0.81-0.89) | 0.86 (0.82-0.90) |
| **Reasons for HIV testing** |  |  |  |
| Just to know HIV status | 70 (62.5) | 1 |  |
| Current/previous HIV risk | 55 (57.3) | 0.97 (0.89-1.05) |  |
| Symptomatic | 216 (53.5) | 0.94 (0.89-1.01) |  |
| **Depression** |  |  |  |
| No depression | 315 (54.6) | 1 |  |
| Major depression | 36 (63.2) | 1.10 (0.97-1.14) |  |
| **Perceived social support** |  |  |  |
| Medium to high | 348 (56.9) | 1 | 1 |
| Low | 6 (20.0) | 0.77 (0.68-0.86) | 0.79 (0.70-0.88) |
| **Expectation of social support after HIV disclosure** |  |  |  |
| Low | 14 (63.6) | 1 |  |
| Medium to high | 340 (54.8) | 0.95 (0.83-1.07) |  |
| **Baseline CD4 count at testing** |  |  |  |
| <350 | 99 (52.7) | 0.97 (0.89-1.06) |  |
| 350-500 | 36 (63.2) | 1.04 (0.94-1.16) |  |
| >500 | 41 (56.9) | 1 |  |
| Missing | 178 (54.8) | 0.99 (0.91-1.07) |  |
| **Initiated on ART** |  |  |  |
| Initiated on ART | 296 (56.9) | 0.94 (0.88-1.00) |  |
| No ART initiation | 58 (47.5) | 1.57 (1.53-1.16) |  |

### High anticipated stigma among newly diagnosed patients

Overall, 55% of the participants were classified as having high anticipated stigma while 45% were classified as having low to medium anticipated stigma (**Fig 1**). Among male participants, 56% had high anticipated stigma compared to 54% of females in the same population. Regarding age, the majority (50.7%) of participants with high anticipated stigma were aged between 30 and 40 years, 35% were aged between 18 and 29 years, and 24% were older than 40 years at the time of enrollment. Regarding marital status, 43% of those who married, 60% of those who were in a cohabiting relationship, 56% of those who were in a non-cohabiting relationship and 54% of those who were single had high anticipated stigma.

**Fig 1.**
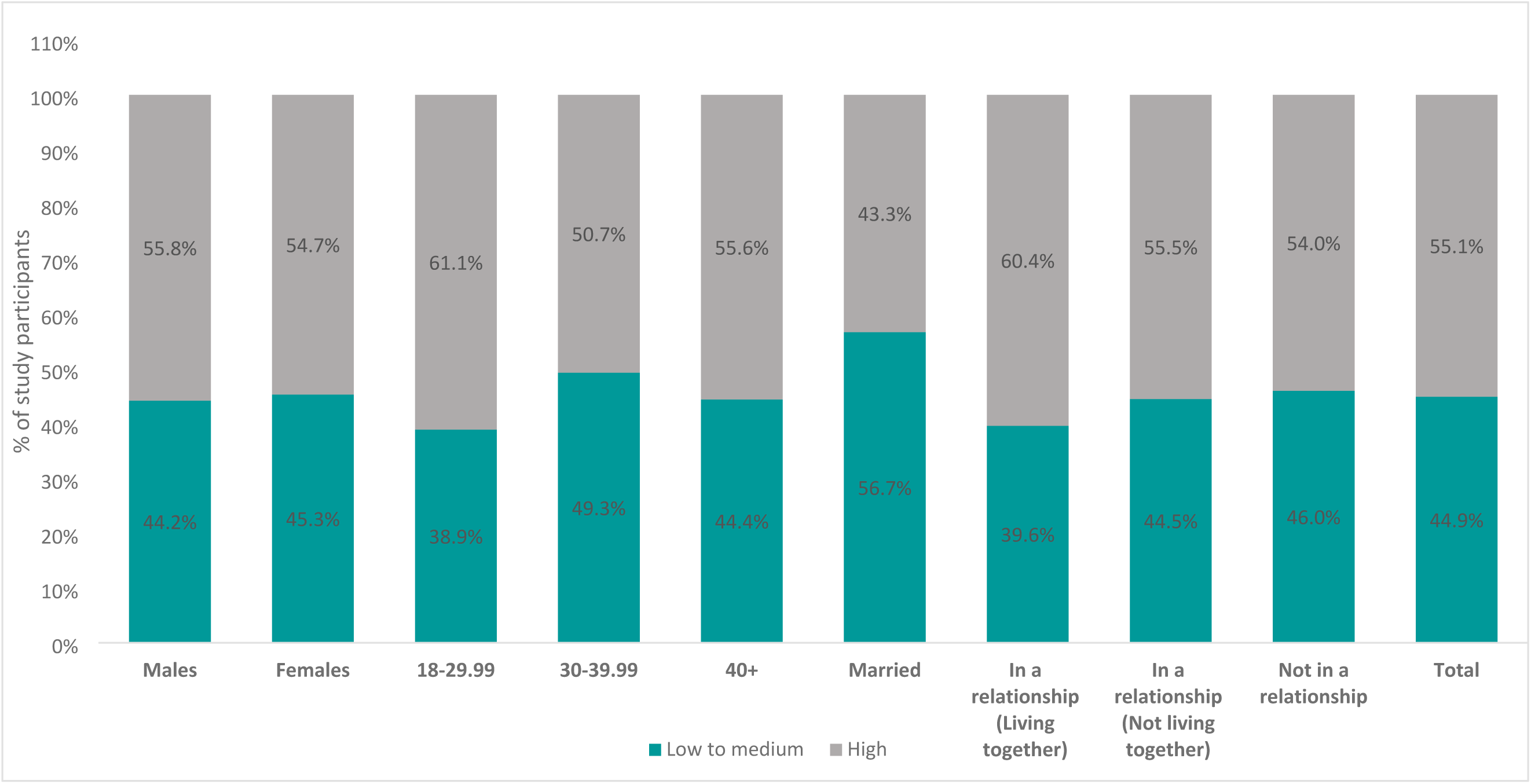
Prevalence of high anticipated HIV stigma

### Correlates of high anticipated HIV stigma among newly diagnosed

After adjusting for demographic patients’ characteristics, unmarried individuals who were in a relationship were more likely to have high anticipated stigma than married participants (aRR 1.10, 95% CI: 1.01-1.18). High anticipated stigma was lower among: older individuals (aRR 0.94 for being 30-39 vs 18-29 years, 95% CI: 0.88-0.99), those having a primary house in another province/rural (aRR 0.82 for primary house in another country vs current house, 95% CI: 0.78-0.87), (aRR 0.83 for primary house in another country vs current house, 95% CI: 0.78-0.88), those living in current homes for ≥5 years (aRR 0.93 for >5 years vs <1 year, 95% CI: 0.88-0.99), those with low ART concerns (aRR 0.86, 95 % CI: 0.82-0.90), and those with low perceived social-support (aRR 0.79 for low vs high, 95 % CI: 0.70-0.88). However, gender, level of education, person to whom they had they had intended to disclose, reason for HIV testing, and level of depression did not show association with anticipated stigma among the newly diagnosed.

## DISCUSSION

The current study measured the prevalence of anticipated stigma among patients newly diagnosed with HIV between October 2017 to August 2018. Our results indicate that over 50% of this population had high anticipated stigma, which is a cause for concern, considering all of these participants had just received HIV counselling. This result indicates that despite the scale of treatment in South Africa and advances to make HIV more manageable, the expectation of social stigma remains a challenge[26].

The findings also showed similarly high anticipated stigma for male and female participants. These findings are consistent with findings from studies in South Africa, which indicated that both males and females had high levels of external stigma [27, 28]. In other settings, however, several studies have shown stigma to be higher in women compared to men [29–31], as women are traditionally held to higher moral standards than men and expected to uphold the moral foundation of society. But other studies did not find gender differences in relation to anticipated stigma[32]. This could be explained by studies that recruit participants from the clinic-based settings where men and women might already be accessing HIV care. These shared experiences may reduce perceived stigma differences by gender. Furthermore, in settings where treatment access and awareness are high, stigma may be declining in both genders, making it harder to detect if there are any differences.

Furthermore, unmarried individuals who were in a relationship reported higher anticipated stigma than those who were married. Previous research has shown that anticipated stigma is significantly associated with non-disclosure of HIV status, particularly among women. This barrier is especially pronounced among women who are not in stable relationships, where fear of rejection or negative consequences can discourage disclosure[33, 34]. While stigmatizing attitudes held by their communities may affect all living with HIV, those who are married or living with a partner may have additional emotional and social support that acts as a barrier to community-level prejudice[35, 36]. However, other studies report that married persons also face anticipated stigma. Their concerns or anxieties include the fear of stigmatisation, the fear of divorce or rejection by their partners, intimate partner violence and the fear that they may be accused of infidelity[37]. Unmarried individuals might worry more about social stigma and be more afraid to disclose because there’s a chance they won’t find future sexual partners, and those with partners may fear being rejected and losing those relationships[9]. The findings are, however, in contrast to previous findings, which reported that single participants reported less stigma compared to their counterparts[38].

Conversely, older participants (30 -39) reported a lower risk of anticipated stigma compared to younger participants, and this finding is consistent with previous findings that normalisation of HIV as a chronic condition, whereby older people compare HIV to other chronic conditions such as hypertension and diabetes[39]. Similarly, those with low ART concerns were less likely to report anticipated stigma, this could be explained by the fact that they have higher factual knowledge of HIV/ART[40]. Knowledge was shown to be associated with reduced concerns as they are more aware of their diagnosis and treatment and generally have a more positive outlook.

Furthermore, those with low perceived social support were also less likely to report anticipated stigma. This could be as a result of them having developed coping mechanisms that help them handle stigma better and potential for alternative sources of validation. The findings are again in contrast to previous research that has shown that the support from friends and family is valuable to counter stigma[41, 42].

### Study limitations

Firstly, we only surveyed HIV positive patients who had engaged in healthcare services; it is possible that some HIV positive people are more likely to report anticipated stigma and would have completely avoided testing, and this could explain the inability to observe some of the previously reported factors. Secondly, we interviewed patients immediately after their post-test counselling and it is possible that some may have, and it is possible that some may not have had sufficient time to adequately process their diagnosis and think about other aspects of their lives.

## CONCLUSIONS

Our findings show that over 50% of adults diagnosed with HIV in the UTT era experienced high anticipated stigma. These findings suggest that stigma remains pervasive and highlight the need to address factors that continue to drive anticipated stigma, to mitigate the potential impact on engagement in HIV care. HIV programs need to be more focused on strategies that directly address stigma upon diagnosis.

## Data Availability

The de-identified data supporting the findings of this study are available from the corresponding author upon reasonable request, subject to ethical approvals and data use agreements in place.

## ACKNOWLEDGEMENTS

We extend our gratitude to the data collection team (Julia Mogwasi, Jan Maobela, Nokukhanya Mhlanga, Vinolia Njinkelanga, Zoleka Luvuno, Nonhlanhla Tshabalala) for their diligent support during the study implementation. Additionally, our sincere thanks go to the City of Johannesburg and the staff of participating clinics for accommodating the study. We also thank patients attending these clinics for their willingness to participate and share their valuable information.

## REFERENCES

1. UNAIDS. UNAIDS 90-90-90: an ambitious treatment target to help end the AIDS epidemic. UNAIDS Geneva, Switzerland; 2014.

2. Nyasulu J, Maposa I. Progress towards 90-90-90 and 95-95-95 strategy implementations and HIV positivity trends in the City of Johannesburg. South African Medical Journal. 2024;114(1):45–9.

3. NDoH. National Policy on HIV Pre-exposure Prophylaxis (PrEP) and Test and Treat (T&T). Pretoria: National Department of Health. 2016.

4. NDOH. Fast tracking implementation of the 90-90-90 strategy for HIV, through implementation of the test and treat (TT) policy and same-day anti-retroviral therapy (ART) initiation for positive patients. Pretoria, South Africa: National Department of Health (NDOH)

5. 2017.

6. Nyongesa MK, Nasambu C, Mapenzi R, Koot HM, Cuijpers P, Newton CR, et al. Psychosocial and mental health challenges faced by emerging adults living with HIV and support systems aiding their positive coping: a qualitative study from the Kenyan coast. BMC Public Health. 2022;22(1):1–20.

7. Geffen N, Cameron E. The deadly hand of denial: Governance and politically-instigated AIDS denialism in South Africa. 2009.

8. Chigwedere P, Seage III GR, Gruskin S, Lee T-H, Essex M. Estimating the lost benefits of antiretroviral drug use in South Africa. JAIDS Journal of Acquired Immune Deficiency Syndromes. 2008;49(4):410–5.

9. Simbayi L, Zuma K, Cloete A, Jooste S, Zimela S, Blose S, et al. The people: Living with HIV stigma index: South Africa 2014: Summary report. 2015.

10. Sineke T, Onoya D, Mokhele I, Cele R, Sharma S, Sigasa P, et al. “I was scared dating… who would take me with my status?”—Living with HIV in the era of UTT and U= U: A qualitative study in Johannesburg, South Africa. PLOS Global Public Health. 2023;3(10):e0000829.

11. Davis A, Pala AN, Nguyen N, Robbins RN, Joska J, Gouse H, et al. Sociodemographic and psychosocial predictors of longitudinal antiretroviral therapy (ART) adherence among first-time ART initiators in Cape Town, South Africa. AIDS care. 2021;33(11):1394–403.

12. Kalichman SC, Kalichman MO. HIV-related stress and life chaos mediate the association between poverty and medication adherence among people living with HIV/AIDS. Journal of clinical psychology in medical settings. 2016;23:420–30.

13. Stern E, Colvin C, Gxabagxaba N, Schutz C, Burton R, Meintjes G. Conceptions of agency and constraint for HIV-positive patients and healthcare workers to support long-term engagement with antiretroviral therapy care in Khayelitsha, South Africa. African Journal of AIDS Research. 2017;16(1):19–29.

14. Earnshaw VA, Smith LR, Chaudoir SR, Amico KR, Copenhaver MM. HIV stigma mechanisms and well-being among PLWH: a test of the HIV stigma framework. AIDS and Behavior. 2013;17:1785–95.

15. Camacho G, Kalichman S, Katner H. Anticipated HIV-related stigma and HIV treatment adherence: the indirect effect of medication concerns. AIDS and Behavior. 2020;24:185–91.

16. Mojola SA, Angotti N, Denardo D, Schatz E, Xavier Gómez Olivé F. The end of AIDS? HIV and the new landscape of illness in rural South Africa. Global Public Health. 2022;17(1):13–25.

17. Viljoen L, Bond VA, Reynolds LJ, Mubekapi-Musadaidzwa C, Baloyi D, Ndubani R, et al. Universal HIV testing and treatment and HIV stigma reduction: a comparative thematic analysis of qualitative data from the HPTN 071 (PopART) trial in South Africa and Zambia. Sociology of Health & Illness. 2021;43(1):167–85. doi: 10.1111/1467-9566.13208.

18. Turan B, Budhwani H, Fazeli PL, Browning WR, Raper JL, Mugavero MJ, et al. How does stigma affect people living with HIV? The mediating roles of internalized and anticipated HIV stigma in the effects of perceived community stigma on health and psychosocial outcomes. AIDS and Behavior. 2017;21:283–91.

19. Bogart LM, Cowgill BO, Kennedy D, Ryan G, Murphy DA, Elijah J, et al. HIV-related stigma among people with HIV and their families: a qualitative analysis. AIDS and Behavior. 2008;12:244–54.

20. MacLean JR, Wetherall K. The association between HIV-stigma and depressive symptoms among people living with HIV/AIDS: A systematic review of studies conducted in South Africa. Journal of affective disorders. 2021;287:125–37.

21. Mall S, Middelkoop K, Mark D, Wood R, Bekker L-G. Changing patterns in HIV/AIDS stigma and uptake of voluntary counselling and testing services: the results of two consecutive community surveys conducted in the Western Cape, South Africa. AIDS care. 2013;25(2):194–201.

22. Pantelic M, Sprague L, Stangl AL. It’s not “all in your head”: critical knowledge gaps on internalized HIV stigma and a call for integrating social and structural conceptualizations. BMC infectious diseases. 2019;19:1–8.

23. Council SANA. National Strategic Plan for HIV, TB and STIs 2023-2028. SANAC website. 2023.

24. Onoya D, Sineke T, Mokhele I, Bor J, Fox MP, Miot J. Understanding the reasons for deferring ART among patients diagnosed under the same-Day-ART policy in Johannesburg, South Africa. AIDS and Behavior. 2021;25(9):2779–92.

25. Berger BE, Ferrans CE, Lashley FR. Measuring stigma in people with HIV: Psychometric assessment of the HIV stigma scale¶. Research in nursing & health. 2001;24(6):518–29.

26. Dos Santos MM, Kruger P, Mellors SE, Wolvaardt G, Van Der Ryst E. An exploratory survey measuring stigma and discrimination experienced by people living with HIV/AIDS in South Africa: the People Living with HIV Stigma Index. BMC public health. 2014;14:1–13.

27. Grossman CI, Stangl AL. Global action to reduce HIV stigma and discrimination. Wiley Online Library; 2013. p. 18881.

28. Ncitakalo N, Mabaso M, Joska J, Simbayi L. Factors associated with external HIV-related stigma and psychological distress among people living with HIV in South Africa. SSM-Population Health. 2021;14:100809.

29. Treves-Kagan S, El Ayadi AM, Pettifor A, MacPhail C, Twine R, Maman S, et al. Gender, HIV Testing and Stigma: The Association of HIV Testing Behaviors and Community-Level and Individual-Level Stigma in Rural South Africa Differ for Men and Women. AIDS and Behavior. 2017;21(9):2579–88. doi: 10.1007/s10461-016-1671-8.

30. Bond V, Levy C, Clay S, Kafuma T, Nyblade L, Bettega N. Kanayaka: “The light is on”: Understanding HIV and AIDS related Stigma in Urban and Rural Zambia. 2003.

31. Loutfy MR, Logie CH, Zhang Y, Blitz SL, Margolese SL, Tharao WE, et al. Gender and ethnicity differences in HIV-related stigma experienced by people living with HIV in Ontario, Canada. PloS one. 2012;7(12):e48168.

32. Nyblade L, Pande R, Mathur S, MacQuarrie K, Kidd R, Banteyerga H, et al. Disentangling HIV and AIDS stigma in Ethiopia, Tanzania and Zambia. 2003.

33. Ataro Z, Mengesha MM, Abrham A, Digaffe T. Gender differences in perceived stigma and coping strategies among people living with HIV/AIDS at jugal hospital, Harar, Ethiopia. Psychology Research and Behavior Management. 2020:1191–200.

34. Ojikutu BO, Pathak S, Srithanaviboonchai K, Limbada M, Friedman R, Li S, et al. Community cultural norms, stigma and disclosure to sexual partners among women living with HIV in Thailand, Brazil and Zambia (HPTN 063). PloS one. 2016;11(5):e0153600.

35. Tam M, Amzel A, Phelps BR. Disclosure of HIV serostatus among pregnant and postpartum women in sub-Saharan Africa: a systematic review. AIDS care. 2015;27(4):436–50.

36. Gutin SA, Ruark A, Darbes LA, Neilands TB, Mkandawire J, Conroy AA. Supportive couple relationships buffer against the harms of HIV stigma on HIV treatment adherence. BMC Public Health. 2023;23(1):1878.

37. Luthuli MQ, John-Langba J. The Moderating Role of HIV Stigma on the Relationship between Perceived Social Support and Antiretroviral Therapy Adherence Self-Efficacy among Adult PLHIV in South Africa. Journal of the International Association of Providers of AIDS Care (JIAPAC). 2024;23:23259582241228743.

38. Bhatia DS, Harrison AD, Kubeka M, Milford C, Kaida A, Bajunirwe F, et al. The role of relationship dynamics and gender inequalities as barriers to HIV-serostatus disclosure: qualitative study among women and men living with HIV in Durban, South Africa. Frontiers in public health. 2017;5:188.

39. Chekole YA, Tarekegn D. HIV-related perceived stigma and associated factors among patients with HIV, Dilla, Ethiopia: a cross-sectional study. Annals of Medicine and Surgery. 2021;71:102921.

40. Emlet CA. A comparison of HIV stigma and disclosure patterns between older and younger adults living with HIV/AIDS. AIDS Patient Care & STDs. 2006;20(5):350–8.

41. Sineke T, Mokhele I, Langa J, Mngoma B, Onoya D. HIV and ART related knowledge among newly diagnosed patients with HIV under the universal-test-and-treat (UTT) policy in Johannesburg, South Africa. AIDS care. 2022;34(5):655–62.

42. Andrews S. Social support as a stress buffer among human immunodeficiency virus-seropositive urban mothers. Holistic Nursing Practice. 1995;10(1):36–43.

43. Galvan FH, Davis EM, Banks D, Bing EG. HIV stigma and social support among African Americans. AIDS patient care and STDs. 2008;22(5):423–36.

